# Estimating the contribution of adults living with HIV to vaccine serotype pneumococcal transmission in Malawi after longstanding infant PCV13 vaccination

**DOI:** 10.64898/2026.11.23.26368808

**Authors:** Deus Thindwa, Lukerensia Mlongoti, Ndaona Mitole, Alice Kusakala, Mercy Khwiya, Thokozani Kayembe, Edwin Lisimba, Joseph Aston Phiri, Lusako Lucky Sibale, Kondwani Charles Jambo

**Author notes:** **Correspondence** Deus Thindwa, Kondwani Jambo, **Address** Deus Thindwa, PhD, Department of Health Data Science, Institute of Population Health, Faculty of Health & Life Sciences, Waterhouse Building, Block F, Liverpool L69 3GF, University of Liverpool.

## Abstract

Despite longstanding infant pneumococcal conjugate vaccine (PCV13), residual vaccine-type (VT) carriage persists in Malawi, where human immunodeficiency virus (HIV) prevalence is high. Adults living with HIV (ALWH) on long-term (>1 year) antiretroviral therapy (ART) show higher pneumococcal carriage and shedding than HIV-uninfected adults, but their contribution to VT transmission remains unclear. We conducted a longitudinal household study in Malawi with weekly nasopharyngeal sampling and fitted a household transmission dynamic model using Markov Chain Monte Carlo methods. We collected 906 samples from 302 participants: younger children (0-5 years), older children (6-17 years) and adults (18+ years) stratified by HIV status and ART duration. VT transmission was more intense with household mixing, with ALWH on long-term ART contributing most to onward transmission to younger children. In contrast, non-vaccine serotype (NVT) transmission was driven mainly by younger children across multiple household pathways. Younger children and ALWH on long-term ART had prolonged carriage, which may have sustained household transmission. VT acquisition was driven mainly by household exposure, whereas NVT acquisition was balanced between household and community sources. Unvaccinated ALWH substantially contribute to household pneumococcal transmission, sustaining residual VT carriage. Optimal pneumococcal vaccination strategies targeting ALWH should be evaluated in high HIV prevalence low-income settings.

## Introduction

*Streptococcus pneumoniae* (pneumococcus) remains a leading cause of morbidity and mortality worldwide, accounting for 300,000–400,000 deaths annually among children under five years of age, with the greatest burden in sub-Saharan Africa ^1,2^. Pneumococcal colonisation of the nasopharynx is a prerequisite for both onward transmission and the development of pneumococcal disease ^3–5^. Understanding the dynamics of pneumococcal carriage and transmission is therefore critical for optimising public health interventions.

The use of pneumococcal conjugate vaccines (PCVs) has substantially reduced vaccine-serotype (VT) carriage and invasive pneumococcal disease (IPD), both directly in vaccinated children and indirectly in unvaccinated older children and adults ^6^. These indirect effects are mediated through disruption of pneumococcal carriage and subsequent person-to-person transmission ^4,7^. However, despite longstanding infant PCV programmes, residual VT carriage and transmission persist in several low-income settings ^8^, where the transmission intensity and epidemiological context differ substantially from those in high-income countries.

Malawi introduced the 13-valent pneumococcal conjugate vaccine (PCV13) into the national immunisation programme in 2011 using a 3+0 schedule (at 6, 10, 14 weeks old) ^9^. Subsequent studies demonstrated reductions in VT carriage and VT IPD among both vaccinated children and unvaccinated older children and adults ^2,7,8,10,11^, alongside a slight increase in non-vaccine serotypes (NVTs) ^11^. Nevertheless, VT carriage has persisted despite vaccine coverage exceeding 90% ^7,8,12^. An earlier modelling study in Blantyre found a local and age-dependent force of infection (FOI) as a determinant of residual VT carriage ^13^. However, the transmission-route profiles on who acquires pneumococci from whom remain incompletely understood.

Children under five years of age are traditionally considered the principal reservoir for pneumococcal transmission because of their high carriage prevalence and prolonged carriage duration ^14,15^. However, the relative importance and directionality of transmission pathways, including the contribution of within-household versus community exposure to the overall local FOI, may shift following a long-standing infant PCV programme ^15–17^. In particular, reductions in VT carriage among vaccinated children may alter the traditionally child-centred transmission network, increasing the relative contribution of alternative transmission pathways and reservoirs. Transmission dynamic modelling provides a robust framework for disentangling transmission pathways and estimating the relative importance of carriage sources to transmission ^4,16,17^.

A potential reservoir of VT transmission includes adults (18+ years) living with HIV (ALWH) on long-term (>1 year) antiretroviral therapy (ART), who are characterised with higher pneumococcal carriage prevalence than HIV-uninfected adults or adults recently initiating on ART (<3 months) ^10^. In addition, ALWH on long-term ART have been shown to shed pneumococci more frequently than HIV-uninfected adults ^18^, suggesting increased potential for onward transmission including residual VT carriage despite longstanding infant PCV13 vaccination. The contribution of ALWH on long-term ART to pneumococcal transmission in the mature PCV13 era remains poorly understood.

Here, we combined the longitudinal household nasopharyngeal carriage surveillance data from Malawi with the transmission dynamic model and Markov Chain Monte Carlo methods to estimate the contribution of ALWH to residual VT pneumococcal transmission in a mature PCV13 era.

## Results

### Baseline characteristics and pneumococcal carriage patterns

At baseline, a total of 302 participants from 67 households were included in the longitudinal household study. NVT carriage was most frequently observed among younger children aged 0–5 years (67.1%) and ALWH receiving long-term ART (61.9%). VT carriage was most frequently observed among older children aged 6–17 years (12.7%). Carriage prevalence varied minimally by sex and household size (Table 1). Across the three weekly sampling visits, VT carriage prevalence remained relatively stable (10.3–11.6%). NVT carriage prevalence ranged from 42.4% to 47.7% across visits. These observations suggested persistent residual VT circulation alongside widespread NVT carriage in this mature infant PCV13 setting (Table 1).

**Table 1.** Baseline characteristics of the study participants during longitudinal nasopharyngeal swabbing household study in Blantyre, Malawi.

| **Number of study participants (N = 302)** | **Uncolonized** | **Vaccine serotypes (VT)<sup>§</sup>** | **Non-vaccine serotypes (NVT)<sup>§</sup>** |
|---|---|---|---|
| Variable | n (%) | n (%) | n (%) |
| Weekly sampling |  |  |  |
| Visit 1 (baseline) | 127 (42.0) | 31 (10.3) | 144 (47.7) |
| Visit 2 | 139 (46.0) | 35 (11.6) | 128 (42.4) |
| Visit 3 | 137 (45.4) | 35 (11.6) | 130 (43.0) |
| Mean age, years (SD)<sup>*</sup> | 23.8 (14.3) | 16.1 (12.9) | 15.7 (13.8) |
| Age/HIV group (years, y)<sup>†</sup> |  |  |  |
| Young children (0-5y) | 16 (21.1) | 9 (11.8) | 51 (67.1) |
| Older children (6-17y) | 30 (38.0) | 10 (12.7) | 39 (49.3) |
| Index Adults (18+y) HIV- | 13 (68.4) | 2 (10.5) | 4 (21.1) |
| Index Adults (18+y) HIV+ ART-S | 14 (51.9) | 2 (7.4) | 11 (40.7) |
| Index Adults (18+y) HIV+ ART-L | 6 (28.6) | 2 (9.5) | 13 (61.9) |
| Household Adults (18+y) | 48 (60.0) | 6 (7.5) | 26 (32.5) |
| Sex |  |  |  |
| Female | 63 (40.4) | 15 (9.6) | 78 (50.0) |
| Male | 64 (43.8) | 16 (11.0) | 66 (45.2) |
| Mean household size (SD)<sup>*</sup> | 5.1 (1.7) | 5.4 (2.1) | 5.1 (1.7) |
| Family size |  |  |  |
| 2 | 9 (45.0) | 2 (10.0) | 9 (45.0) |
| 3-5 | 77 (40.5) | 19 (10.0) | 94 (49.5) |
| 6+ | 41 (44.6) | 10 (10.8) | 41 (44.6) |

<sup>§</sup> Pneumococcal conjugate vaccine (PCV13) serotypes (VT) and non-PCV13 serotypes (NVT)
<sup>\*</sup> SD - Standard deviation
<sup>†</sup> Human immunodeficiency virus (HIV) infection status in adults
<sup>†</sup> Antiretroviral therapy (ART)
<sup>†</sup> HIV- (without HIV infection), HIV+ ART-S (with HIV and shorter duration of ART use of <3 months), HIV+ ART-L (with HIV and longer duration of ART use of >1 year)

### ALWH are important contributors to household residual VT transmission

Household VT carriage was transmitted through distinct routes, with ALWH on long-term ART being important contributors to onward transmission to younger children (posterior median transmission rate β = 1.25 weekly transmission events; 95% credible interval [95%CrI]: 0.34–2.62) (Fig. 1A). This pathway accounted for an estimated 38.8% of the household VT FOI experienced by younger children (Table S1). Other notable pathways included household VT transmission from older children to younger children (β = 1.08, 95%CrI: 0.23–2.38) (Fig. 1A), accounting for 33.1% of household VT FOI in younger children (Table S1), and household VT transmission from older children to HIV-uninfected adults (β = 1.18, 95%CrI: 0.28–2.59) (Fig. 1A), accounting for 35.7% of household VT FOI in HIV-uninfected adults (Table S1). Overall, the onward household VT transmission occurred through a relatively limited number of pathways involving ALWH on long-term ART and older children, with considerable uncertainty due to limited sample size.

**Figure 1.**
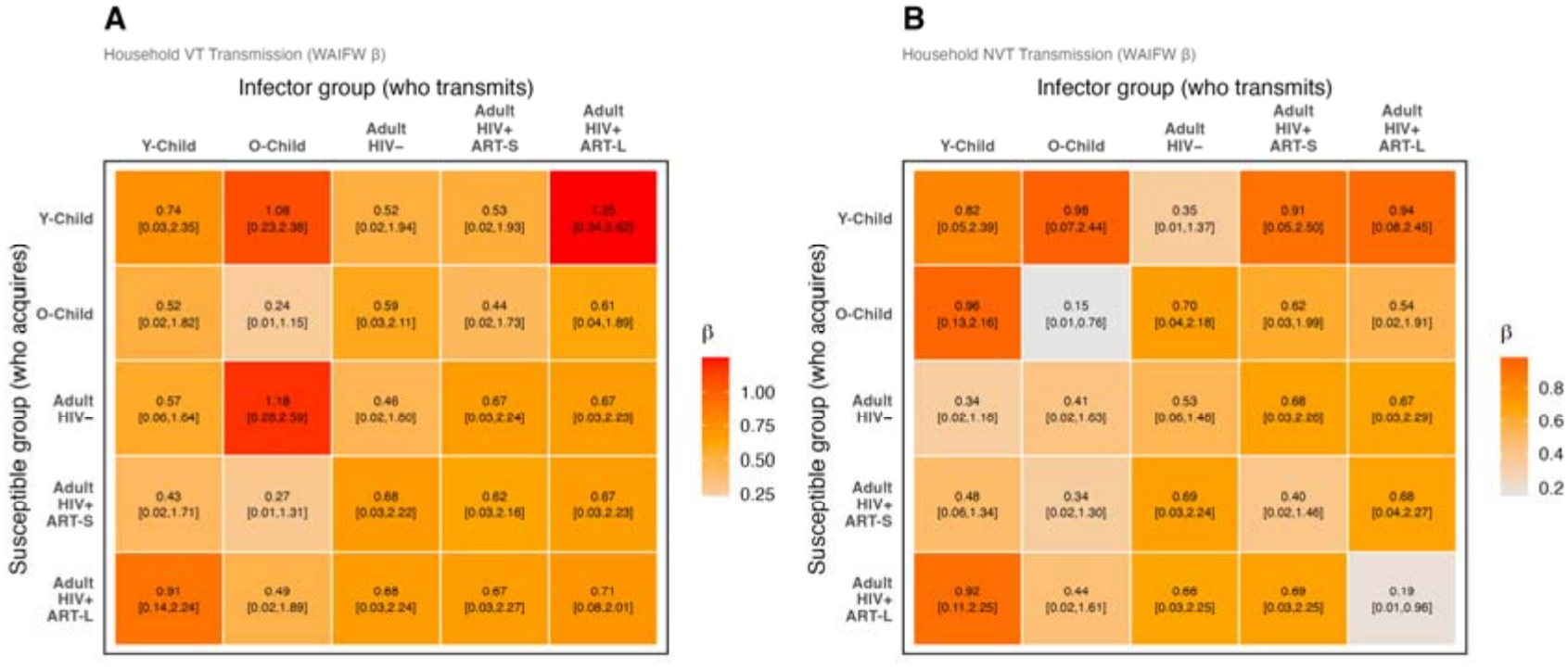
Heatmaps of who acquires infection from whom (WAIFW) matrices representing routes of weekly pneumococcal transmission events. The estimates in each cell show the posterior median and 95% credible intervals (CrI) obtained by fitting a household pneumococcal transmission model to longitudinal pneumococcal swabbing data, and sampling from the parameter posterior distribution using Hamiltonian Monte Carlo (HMC) No-U-Turn Sampler (NUTS). The heatmaps show weekly transmission events from age/HIV group to another age/HIV group for (A) vaccine serotypes (VT) and (B) non-vaccine serotypes (NVT). Considered age/HIV groups include younger children 0-5 years-old (Y-Child), older children 6-17 years-old (O-Child), adults 18+ years-old without HIV (Adult HIV-), adults 18+ years-old with HIV and shorter duration (<3 months) of ART use (Adult HIV+ ART-S), and adults 18+ years-old with HIV and prolonged duration (>1 year) of antiretroviral (ART) use (Adult HIV+ ART-L).

Household exposure was a major source of VT acquisition, exceeding 50% of the total VT exposure for each age/HIV group except for older children whose community exposure of VT accounted for 59.7% (95%CrI: 34.3-82.1) (Fig 2A, Fig 2B, Table S2). Compared to community, household VT infection pressure was most pronounced among HIV-uninfected adults (86.6%, 95%CrI: 51.8–99.4). Across age/HIV groups, the posterior distribution of household VT exposure was largely centred between 50% and 85% (Fig. 2C, Table S2). Together, these findings suggest that in this mature PCV13 setting, the residual VT transmission may be sustained predominantly through household contact networks, particularly involving ALWH on long-term ART and older children, with the latter acquiring VT mostly from the community.

**Figure 2.**
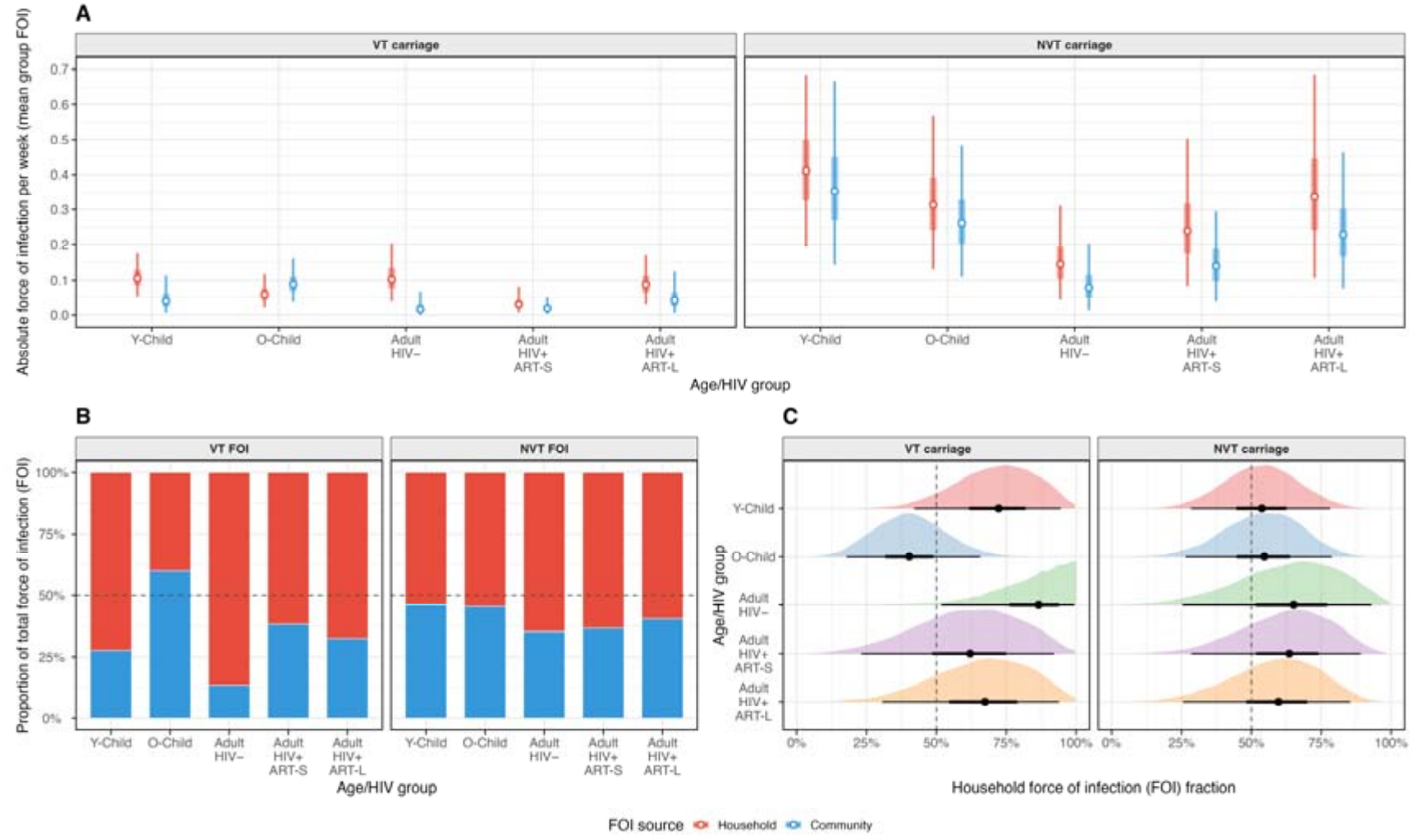
Decomposition of the force of infection (FOI) into household and community sources across age/HIV groups for vaccine serotype (VT) and non-vaccine serotype (NVT) pneumococcal carriage. (A) Posterior median absolute FOI per week with 50% (thick bar) and 95% (thin bar) credible intervals (CrI). (B) Proportional attribution of total FOI to household (red) and community (blue) sources where the posterior distribution of the community FOI is scaled by the community carriage prevalence. (C) Half-eye distribution of the full posterior uncertainty of the household FOI fraction where a point denotes the median, and the horizontal bars denote 50% (thick) and 95% (thin) CrI, and if the 95% CrI spans both sides of 50%, the data cannot distinguish whether household or community is the dominant source for that group. The considered age/HIV groups include younger children 0-5 years-old (Y-Child), older children 6-17 years-old (O-Child), adults 18+ years-old without HIV (Adult HIV-), adults 18+ years-old with HIV and shorter duration (<3 months) of antiretroviral (ART) use (Adult HIV+ ART-S), and adults 18+ years-old with HIV and prolonged duration (>1 year) of ART use (Adult HIV+ ART-L).

### Household NVT transmission involves multiple pathways

Household NVT transmission propagated through multiple potential pathways involving younger children, older children, and ALWH on long-term ART (Fig. 1B, Table S1). Notable estimated transmission routes included those between younger children (β = 0.82 weekly transmission events; 95%CrI: 0.05–2.39), from younger children to older children (β = 0.96, 95%CrI: 0.13–2.16), and from younger children to ALWH on long-term ART (β = 0.92, 95%CrI: 0.11–2.25) (Fig 1B). These pathways accounted for 15.3%, 63.0%, and 68.9% of the household NVT exposure experienced by younger children, older children, and ALWH on long-term ART, respectively (Table S1). Additional estimated pathways included transmission from older children to younger children (β = 0.98, 95%CrI: 0.07–2.44), accounting for 35.1% of the household NVT exposure experienced by younger children, and transmission from ALWH on long-term ART to younger children (β = 0.94, 95%CrI: 0.08–2.45), accounting for 60.6% of the household NVT exposure on younger children (Fig 1B, Table S1). Overall, despite the overlapping uncertainty between pathway estimates, NVT propagation appeared to involve multiple substantial transmission pathways than VT transmission.

In contrast to VT exposure, NVT infection pressure appeared more evenly distributed between household and community sources for each age/HIV group (Fig 2A, Fig 2B, Table S2). The contribution of the household NVT exposure to the total FOI ranged from posterior median 53.7% to 65.1%, moderately higher than that of the community NVT exposure (34.9% to 46.3%). The age/HIV-specific posterior fractions of the household NVT exposure were generally centred compared to those estimated for household VT exposure, highlighting the substantial role of community exposure in driving NVT transmission (Fig. 2C, Table S2). Nevertheless, wider uncertainties were associated with these estimates across age/HIV groups. Together, these findings suggest that NVT circulation in this mature infant PCV13 setting may be sustained by both household and community sources.

### ALWH on long-term ART and younger children largely exhibited prolonged carriage duration

Carriage clearance rates varied across age and HIV groups for both VT and NVT pneumococci (Fig. 3A, Table S3). Younger children exhibited the slowest estimated carriage clearance rates for both serotype groups, corresponding to the longest carriage durations. For younger children, the posterior median weekly clearance rates were 0.30 (95%CrI: 0.07–0.72) for VT carriage and 0.20 (95%CrI: 0.10–0.36) for NVT carriage, corresponding to estimated median carriage durations of 3.4 weeks and 5.0, respectively (Fig. 4, Table S3). ALWH on long-term ART also demonstrated relatively prolonged carriage, particularly for VT pneumococci. Among this group, the posterior median weekly clearance rate for VT carriage was 0.28 (95%CrI: 0.04–0.81) whereas NVT carriage was faster at 0.71 (95%CrI: 0.37–1.22) (Fig. 4, Table S3). Overall, these findings suggest that younger children and ALWH on long-term ART experience prolonged pneumococcal carriage, which likely facilitates onward household transmission.

**Figure 3.**
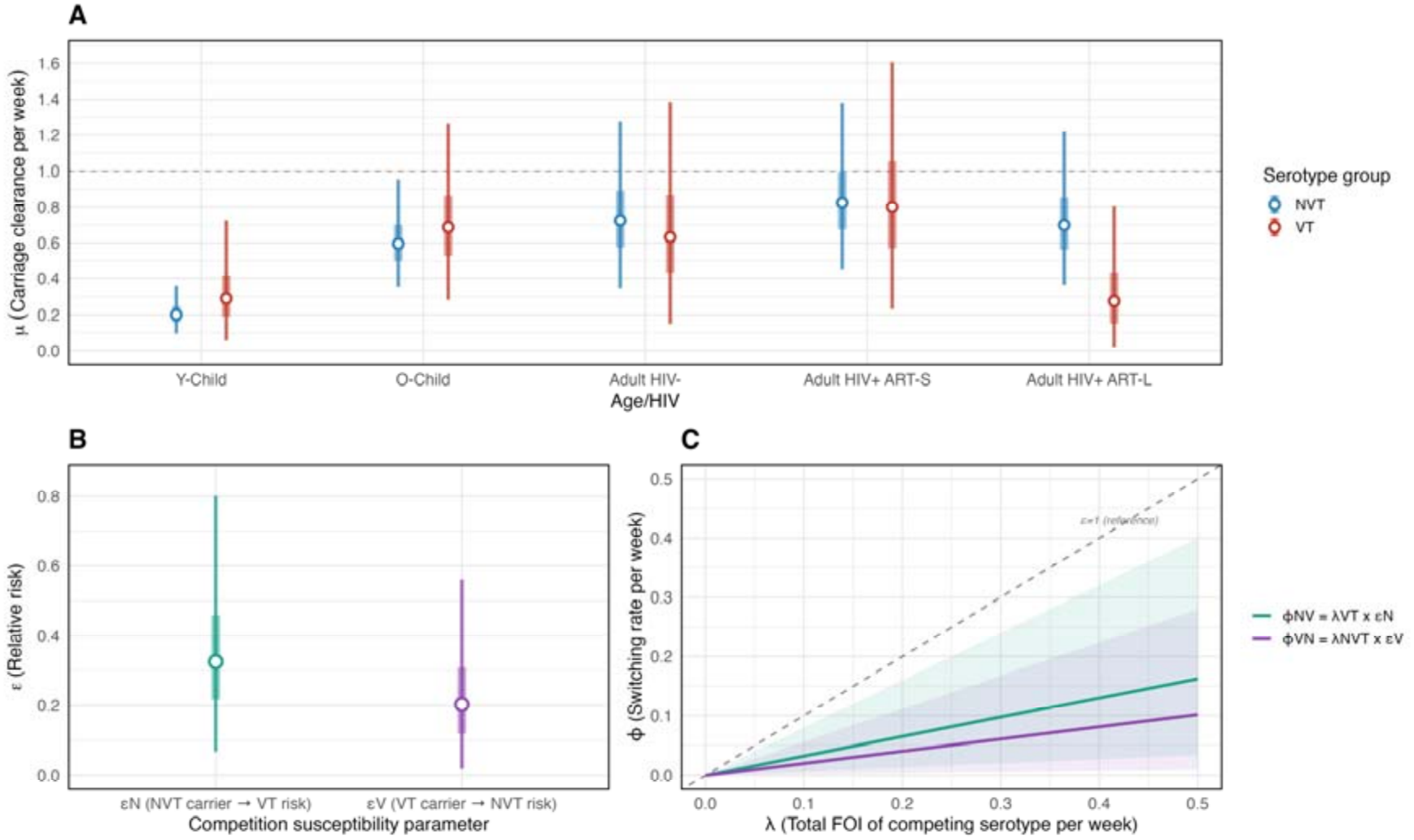
Pneumococcal carriage clearance and competition stratified by vaccine serotype (VT) and non-vaccine serotype group (NVT). All estimates of the competition and weekly pneumococcal carriage clearance refer to the posterior median (point) and 50%, 95% credible intervals (CrI) (thick and thin bars). (A) Weekly carriage clearance of VT and NVT stratified by age/HIV group. (B) The relative risk of acquiring VT while carrying NVT and the relative risk of acquiring NVT while carrying VT. (C) The weekly competition rate against weekly FOI of the competing serotype, with dashed line indicating a relative risk of competition of 1 (reference). The considered age/HIV groups include younger children 0-5 years-old (Y-Child), older children 6-17 years-old (O-Child), adults 18+ years-old without HIV (Adult HIV-), adults 18+ years-old with HIV and shorter duration (<3 months) of antiretroviral (ART) use (Adult HIV+ ART-S), and adults 18+ years-old with HIV and prolonged duration (>1 year) of ART use (Adult HIV+ ART-L).

**Figure 4.**
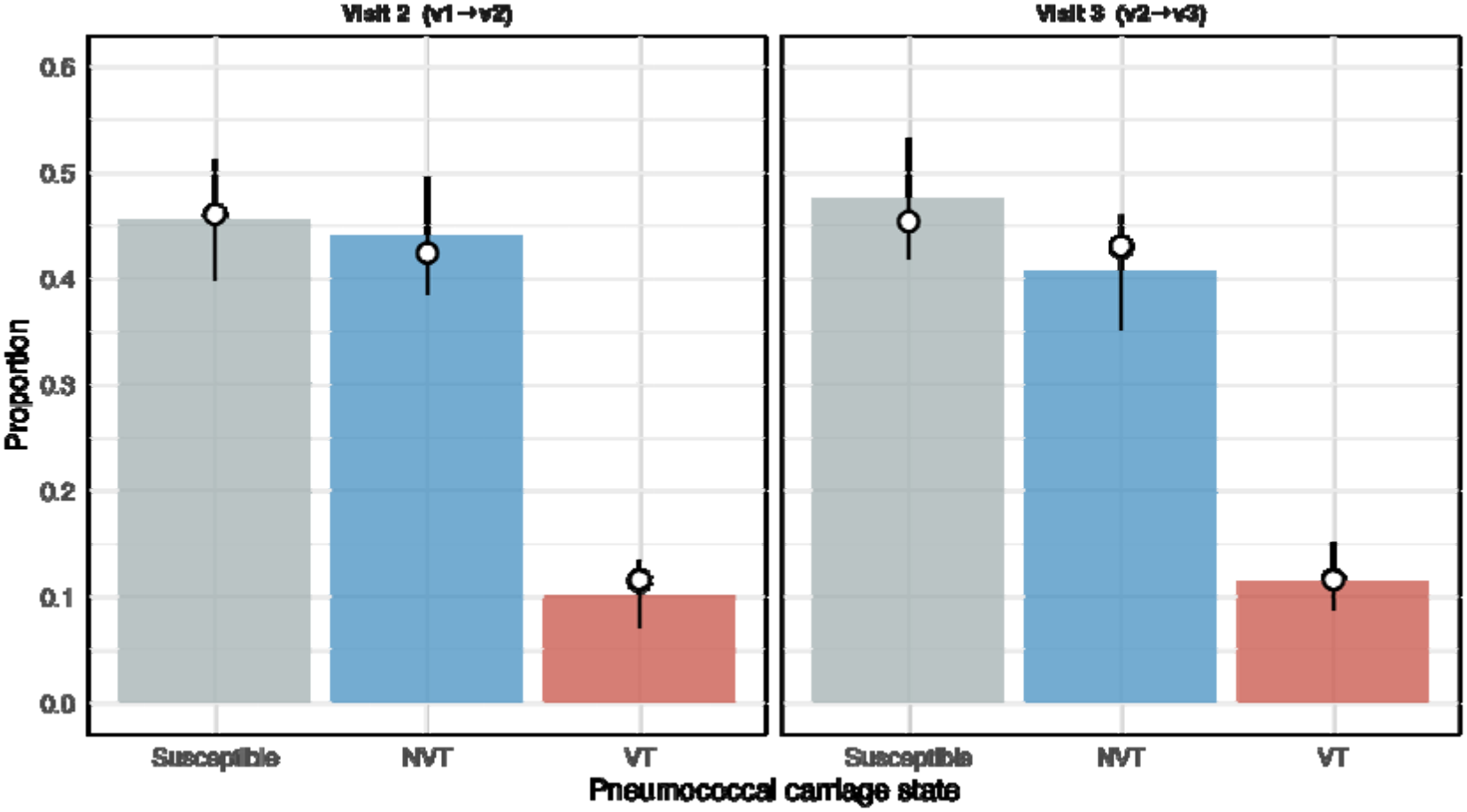
Posterior predictive checks of the simulated or newly replicated datasets based on the fitted model parameters, comparing the predicted median and 95% credible intervals of the replicated dataset to the proportion of the observed carriage dataset during the two study visits. The black circles show the observed carriage proportions. The bar height shows the posterior median and the black vertical lines through the black circle denote the 95% posterior predictive intervals of the estimated proportion. The posterior predictions were based on 16,000 model-simulations after fitting the model to the household swabbing data of 302 members.

### Model validation and sensitivity analyses

The transmission model well fitted to the data and demonstrated good convergence and parameter posterior sampling, with no divergent transitions, *Rhat* equal to 1.00 for all parameters, and minimum bulk effective sample sizes exceeding 7,800. The posterior predictive intervals showed that the fitted model adequately reproduced the observed carriage-state distributions across sequential follow-up visits (Fig. 4). At visit 2, the predicted posterior median proportions were 0.46 (95%CrI: 0.40–0.51) for non-carriers, 0.44 (95%CrI: 0.38–0.50) for NVT carriers, and 0.10 (95%CrI: 0.07–0.14) for VT carriers, compared with observed proportions of 0.46, 0.42, and 0.12, respectively. Similarly, at visit 3, the predicted posterior median proportions were 0.48 (95%CrI: 0.42–0.53) for non-carriers, 0.41 (95%CrI: 0.35–0.46) for NVT carriers, and 0.12 (95%CrI: 0.09–0.15) for VT carriers, compared with observed proportions of 0.45, 0.43, and 0.12, respectively. Additional assessment of model fitting included parameter trace and density plots shown in the supplement (Fig. S1–S5).

While the primary analysis considered household adults with unknown HIV/ART status to have similar HIV/ART status as the adult index, the sensitivity analyses evaluating three alternative assumptions regarding unknown HIV/ART status among adult household members (all household adults being ALWH on long-term ART, ALWH on short-term ART or HIV-uninfected) produced transmission pathways broadly consistent with the primary analysis (Fig. S6, Fig. S7). In particular, the estimated larger contribution of ALWH on long-term ART to household VT transmission to younger children remained robust across alternative assumptions. Likewise, an important role of younger children in multiple household NVT transmission pathways remained consistent across alternative assumptions. Household composition did not affect the contribution of household FOI to the total exposure. Standardising household sizes to the cohort median household size yielded FOI estimates similar to the primary analysis across age/HIV groups (Fig. S8). Overall, these findings support the robustness of the estimated transmission patterns in the primary analysis, despite the overlapping or considerable uncertainties.

## Discussion

Despite longstanding infant PCV13 vaccination and substantial reductions in VT carriage, residual VT circulation persists in Malawi, and the transmission pathways sustaining this persistence remain incompletely understood. In this study, we used a validated household pneumococcal transmission model implemented within a Bayesian inference framework to investigate the household and community transmission patterns in a mature infant PCV13 setting. We found that ALWH on long-term ART contributed notably to onward VT transmission to younger children, highlighting a potentially important non-traditional transmission pathway in the post-PCV era. In contrast, NVT carriage appeared to circulate through multiple household pathways involving younger children, older children, and ALWH on long-term ART, consistent with the established role of children as important pneumococcal reservoirs ^8,14^. Both adults on long-term ART and younger children exhibited prolonged carriage duration, supporting their role for sustaining household transmission. VT acquisition was primarily associated with within-household exposure, whereas NVT acquisition appeared more evenly balanced between household and community sources. Together, these findings demonstrate structured and heterogeneous pneumococcal transmission patterns in a mature infant PCV programme in a low-income setting, with potential implications for extended-valency vaccines and targeted strategies to reduce residual pneumococcal circulation and disease risk.

Before PCV10 introduction in Kenya, school-age children were reported to contribute substantially to pneumococcal transmission ^4^. Consistent with this, we found that older children frequently contributed to VT transmission to younger children, although the estimated intensity of this pathway appeared lower than that observed among adults on long-term ART. The prominent VT transmission pathway from adults on long-term ART to younger children may reflect caregiver roles characterised by frequent, prolonged close physical contacts within the households ^19,20^. These findings suggest that residual VT circulation in this mature infant PCV13 setting may persist through concentrated household transmission pathways despite longstanding infant vaccination. Our findings further support previous modelling studies showing that pneumococcal acquisition is driven predominantly by within-household rather than community exposure ^16,17,21^. Overall, household exposure accounted for the majority of VT acquisition in this population, although these estimates were associated with considerable uncertainty, as reflected by the wide credible intervals.

In contrast, studies of mother–child pairs in South Africa and daycare cohorts in Finland reported higher rates of pneumococcal acquisition in community and daycare settings, respectively, than in households ^20,22^. However, given the relatively limited use of daycare centres in Malawi, it is unlikely that such settings contribute substantially to pneumococcal transmission relative to household exposure. With the growing urbanisation in low-income settings, it will still be important in the future studies to include and understand the role of additional settings, such as daycare centres, in driving pneumococcus and other respiratory pathogens.

The prolonged duration of NVT carriage among younger children and VT carriage among adults on long-term ART suggests that these groups may serve as important reservoirs for their respective serotypes and contribute to sustained household transmission. Our model recapitulated our previous findings ^10^, demonstrating faster carriage clearance among adults on short-term ART and HIV-negative adults compared with adults on long-term ART, with relatively lower estimated transmission intensity from these groups to others. These findings have potential policy implications; Younger children and adults on long-term ART, who appeared to sustain and transmit pneumococci more frequent within-household, may benefit the most from household-level interventions, including targeted vaccination approaches using PCV13 or extended-valency PCVs. Conversely, older children, who appeared to experience greater VT exposure from community sources, may benefit more from broader population-level strategies to reduce pneumococcal carriage, including catch-up vaccination campaigns. With the indirect protection conferred by PCVs, both targeted and population-wide PCV approaches are likely to also benefit other groups that are not directly reached by vaccination.

Previously, our work in this setting focused on characterising the risk factors and dynamics of pneumococcal acquisition and clearance, as well as elucidating the immunological mechanisms underlying persistent carriage among ALWH on long-term ART ^11,18,23^. The present study adds to the growing body of evidence showing that adults on long-term ART are an underrecognised contributor to residual pneumococcal transmission to younger children in high-burden settings. This is consistent with our previous findings demonstrating increased pneumococcal shedding among ALWH on long-term ART compared with HIV-uninfected adults, supporting the biological plausibility of enhanced onward transmission from this group ^18^.

Nevertheless, several limitations should be considered. First, given the limited sample size, several pathway-specific transmission estimates were associated with substantial uncertainty, although the directions of inferences were robust. Second, the model did not account for dual carriage states, and serotyping was performed using single-colony isolates, which may have underestimated co-colonisation and concurrent serotype carriage. Aggregation of individual serotypes into VT and NVT groups simplified the underlying ecological heterogeneity and may have obscured serotype-specific transmission patterns. Despite these limitations, the consistency of the inferred transmission patterns across multiple complementary analyses, including sensitivity analyses and posterior predictive validation, supports the robustness of the principal findings.

In conclusion, alongside children, ALWH receiving long-term ART may contribute importantly to the residual pneumococcal transmission within households. These findings support further evaluation of targeted vaccination strategies that include ALWH in high-burden settings such as Malawi to further reduce pneumococcal carriage, onward transmission, and disease risk.

## Methods

### Study design and setting

This study was conducted in Blantyre, Malawi, between 17^th^ September 2021 and 11^th^ December 2023 as part of the NASOMUNE study ^10,18,23^. Adult community participants (≥18 years; index participants) were recruited using random community-based sampling across multiple urban communities. The cohort included HIV-uninfected adults, adults living with HIV (ALWH) who had received antiretroviral therapy (ART) for a short-term of <3 months (ART-S), and ALWH who had received ART for a long-term of >1 year (ART-L). Index participants were followed longitudinally with weekly nasopharyngeal (NP) swab collection over 17 weeks. A randomly selected subset of consenting index participants was subsequently enrolled into a household transmission sub-study. Thus, each household included at least one adult with known HIV and ART status. Households were defined as individuals co-residing within the same dwelling, and all consenting household members were followed with weekly NP swab collection over three weeks. The household transmission cohort comprised 302 participants from 67 households. Participants were categorised into five age and HIV/ART groups for the transmission modelling analyses: younger children (0–5 years), older children (6–17 years), HIV-uninfected adults, ALWH on ART-S, and ALWH on ART-L.

### Inclusion and exclusion criteria

Adults aged ≥18 years residing within the study catchment area were eligible for enrolment as index participants. Participants were recruited into one of the three predefined groups based on HIV and ART status: HIV-uninfected adults, ALWH on ART-S, and ALWH on ART-L. HIV and ART status for the index participants were confirmed using clinical records and a double rapid-test algorithm ^24^. For the household transmission sub-study, all consenting or assenting household members residing within the same dwelling as the index participant were eligible for inclusion. Household members without available NP swab data were excluded from the transmission analyses.

### Pneumococcal carriage detection and serotyping

Nasopharyngeal swabs were collected in accordance with World Health Organization (WHO) guidelines and processed using standard microbiological methods ^25^. Pneumococcal isolates were identified by culture and serotyped using latex agglutination performed on a single colony isolate to assign serotype or serogroup ^26^. Serotypes included in the 13-valent pneumococcal conjugate vaccine (PCV13) were classified as vaccine-type (VT), while all other serotypes were classified as non-vaccine-type (NVT).

### Pneumococcal household transmission model

Household pneumococcal carriage dynamics were modelled using a three-state continuous-time Markov chain (CTMC) framework under a Susceptible–Infected–Susceptible (SIS) structure at the individual level. At each sampling visit, individuals occupied one of the three mutually exclusive carriage states: uncolonised, colonised with VT, or colonised with NVT. With three sequential swabbing visits, the study generated two consecutive carriage transitions per participant (visit 1→2 and visit 2→3). To account for heterogeneity in susceptibility, transmissibility, and carriage duration, participants were stratified into five groups described above: younger children, older children, HIV-uninfected adults, ALWH ART-S, and ALWH ART-L. Each household included at least one adult with known HIV and ART status.

We develop a model that is a simplification of the complex dynamics of pneumococcal household transmission. With the SIS model structure, an individual can move from being uncolonized to become a VT carrier or NVT carrier after experiencing age/HIV- and serotype group-specific FOI. An individual can move back from a carrier state to uncolonized state over a time interval after clearing VT carriage or NVT carriage. VT carriers have a ‘change of serotype’ rate to switch to become NVT carriers without a clearance episode. Likewise, NVT carriers have a ‘change of serotype’ rate to switch to become VT carriers without a clearance episode (Text S1).

The total FOI was the sum of within-household FOI and community (background) FOI (Equation). Within-household FOI was the function of the transmission rate matrix, the number of carriers in the household and household size, while the community FOI was scaled by the cross-sectional population prevalence. The transmission rate matrix, separate for VT and NVT, was modelled using Who Acquires Infection From Whom (WAIFW) parameterisation, capturing the directional transmission pathways between age/HIV groups. Direct serotype switching between VT and NVT carriage, without a clearance episode, was included to capture the ecological competition between VT and NVT. Clearance rates for VT and NVT were estimated for each age/HIV group to account for the role of each group’s immunity in modulating carriage duration. Additional transmission parameters were derived from the fitted parameter posterior estimates which included the decomposed forces of infection for household and community, and rates of serotype group switching between VT and NVT. Details of the transmission model, parameter definition, and inference are in the supplement (Text S1).

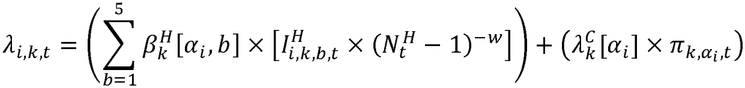

Where *λ*_*i,k,t*_ is force of infection of serogroup *k* at time *t* on an individual i in age/HIV group *α*_*i*_; 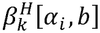 is the household WAIFW transmission rate from an infector in age/HIV group *b* to susceptible in age/HIV group *αi* for serotype group *k* per week. 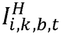 is the count of the serotype group-*k* carriers in age/HIV group *b* in household *H* at visit *t*, excluding the individual *i* themselves. 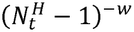 is the total number of other household members at visit *t*, where *w* is the density correlation factor controlling the extent of density-dependency. 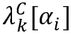 is the community (background) FOI on age/HIV group αi and serotype group *k*. 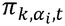 is the observed cross-sectional community prevalence of serotype group *k* in age/HIV group *α*_*i*_ at visit t that scales the community FOI. We assumed the density scaling factor of *w*=1 based on previous studies^16,17^, such that the forces of infection are proportional to the fraction of household members who are carriers.

All model parameters were jointly estimated within a Bayesian inference framework using Hamiltonian Monte Carlo with the No-U-Turn Sampler (NUTS) as implemented in Stan v2.38 ^27^, via RStan interface ^28^. Weakly informative prior distributions were specified for all model parameters (Text S1). Sampling was conducted using four independent Markov chains with 1,000 warm-up iterations and 4,000 post-warm-up iterations per chain, yielding 16,000 posterior samples. Model convergence was assessed using the scale reduction statistic (*Rhat*<1.01), effective sample sizes, divergent transition diagnostics, and energy Bayesian fraction of missing information (E-BFMI). Posterior predictive checks compared observed carriage-state distributions with simulated posterior predictive carriage distributions. Parameter estimates were presented as posterior medians with corresponding 50% or 95% credible intervals (CrIs). All analyses were performed in R v4.5.3 and RStudio v2024.04.0. The mathematical details of the model structure, likelihood formulation, parameterisation, and prior distributions are provided in the Supplement (Text S1).

### Sensitivity analyses

In the primary analysis, adult household members with unknown HIV/ART status were assumed to have a similar HIV/ART status as the household index participant in order to retain the full analytical sample. To assess the robustness of this assumption, sensitivity analyses were conducted where adult household members with unknown HIV/ART status were assumed to be: (a) ALWH on long-term ART, (b) ALWH on short-term ART, or (c) HIV-uninfected. The impact of these assumptions on the estimated within-household transmission rates was evaluated. We also assessed the effect of the household size or composition on the household and community share of the FOI.

## Data Availability

The original data that informed this modelling work contains sensitive clinical information about individual adult HIV infection status and thus is available upon reasonable request to the authors. However, fitted model objects including full posterior data of the main and sensitivity analyses are accessible via Github https://github.com/deusthindwa/spn.hhtransmission.dynamic.model.malawi.

## Ethical approval

Ethical approval for this study was obtained from the Malawi National Health Sciences Research Ethics Committee (NHSRC; 21/24/2680) and the Liverpool School of Tropical Medicine Research Ethics Committee (LSTMREC; 21-035). Written informed consent, and assent where appropriate, was obtained from each parent and/or legal guardian for study participation. All the methods for conducting this study were performed in accordance with the relevant guidelines and regulations as in the approved study protocol by the ethics committees. In addition, all methods for analysing the study data were performed in accordance to relevant guidelines for using infectious disease models for public health research ^29^.

## Acknowledgements

The authors thank all community study participants, and the study staff for their support and co-operation during the study. This work was supported by an African Research Leader (ARL) award [MR/T008822/1] to KCJ. This ARL award is jointly funded by the UK Medical Research Council (MRC) and the UK Foreign Commonwealth and Development Office (FCDO) under the MRC/FCDO Concordat agreement and is also part of the EDCTP2 programme supported by the European Union. A Wellcome Strategic award number 206545/Z/17/Z supports MLW. DT is supported by the African Research Excellence Fund (AREF) Research Development Fellowship (AREF-335-THIN-F-C1114). The funders were not involved in the design of the study; in the collection, analysis, and interpretation of the data; and in writing the manuscript. The findings and conclusions in this report are those of the authors and do not necessarily represent the official position of funders.

## Author contributions

D.T and K.C.J wrote the original draft of the manuscript. D.T curated and analysed the data. D.T, L.M, J.P, L.S and K.C.J revised and edited the manuscript. D.T, J.P, L.S and K.C.J conceptualised the study. D.T, J.P, L.S and K.C.J. validated the study. L.M. and J.P. managed the study data. L.M, D.T, J.P, L.S and K.C.J developed the methodology. L.M conducted the investigation. N.M, A.K and J.P coordinated the study. All authors read, reviewed and approved the submitted version of the manuscript.

## Data availability

### Code availability

Stan/R scripts to reproduce the results reported in this paper are publicly and freely accessible from Github repository: https://github.com/deusthindwa/spn.hhtransmission.dynamic.model.malawi.

### Corresponding authors

Correspondence to Deus Thindwa or Kondwani Jambo

### Declaration of generative AI use

Generative AI was not used in manuscript preparation.

