## Supplementary for "Estimating the contribution of adults living with HIV to vaccine serotype pneumococcal transmission in Malawi after longstanding infant PCV13 vaccination"

### Correspondence

Table of contents

| | |
|---|---|
| Supplementary Table 1 | Attributions of household pneumococcal forces of infections from the infector Age/HIV group to the susceptible Age/HIV group |
| Supplementary Table 2 | Household and community pneumococcal absolute and share of forces of infections on each Age/HIV group |
| Supplementary Table 3 | Age/HIV stratified pneumococcal carriage duration, force of infection, and serotype competition relative risk |
| Supplementary Figure 1 | Trace plots and posterior distribution of vaccine serotype transmission parameters from ‘who acquires infection from who’ (WAIFW) matrices |
| Supplementary Figure 2 | Trace plots and posterior distribution of non-vaccine serotype transmission parameters from ‘who acquires infection from who’ (WAIFW) matrices |
| Supplementary Figure 3 | Trace plots and posterior distribution of vaccine and non-vaccine serotype force of infection parameters by age/HIV groups |
| Supplementary Figure 4 | Trace plots and posterior distribution of vaccine and non-vaccine serotype carriage clearance parameters by age/HIV groups |
| Supplementary Figure 5 | Trace plots and posterior distribution of serotype competition relative risk parameters |
| Supplementary Figure 6 | Heatmaps of posterior estimates of ‘who acquires infection (vaccine serotypes) from who’ (WAIFW) under different assumptions of unknown HIV adult status |
| Supplementary Figure 7 | Heatmaps of posterior estimates of ‘who acquires infection (non-vaccine serotypes) from who’ (WAIFW) under different assumptions of unknown HIV adult status |
| Supplementary Figure 8 | Effect of standardized force of infection (FOI) on household vs community share of FOI |
| Supplementary Text 1 | Details of the household transmission modelling |

| Supplementary Table 1. Attributions of household pneumococcal forces of infections from the infector Age/HIV group to the susceptible Age/HIV group and their estimates of posterior median | | | | | | |
|---|---|---|---|---|---|---|
| | | Infector age/HIV group | | | | |
| | | Young children (0-5 years-old) | Older children (6-17 years-old) | Adults (18+ years-old) HIV- | Adults (18+ years-old) HIV+ ART-S | Adults (18+ years-old) HIV+ ART-L |
| **VT** | | | | | | |
| Susceptible age/HIV group | Young children (0-5 years-old) | 0.00749 | 0.03254 | 0.01547 | 0.00464 | 0.03812 |
| | Older children (6-17 years-old) | 0.01059 | 0.00926 | 0.00161 | 0.00995 | 0.01863 |
| | Adults (18+ years-old) HIV- | 0.04643 | 0.03430 | 0.01523 | 0.00000 | 0.00000 |
| | Adults (18+ years-old) HIV+ ART-S | 0.00760 | 0.01273 | 0.00000 | 0.00600 | 0.00000 |
| | Adults (18+ years-old) HIV+ ART-L | 0.03777 | 0.00765 | 0.00000 | 0.00000 | 0.03607 |
| **NVT** | | | | | | |
| Susceptible age/HIV group | Young children (0-5 years-old) | 0.05806 | 0.13323 | 0.01508 | 0.07947 | 0.09375 |
| | Older children (6-17 years-old) | 0.18243 | 0.01668 | 0.01550 | 0.04211 | 0.03296 |
| | Adults (18+ years-old) HIV- | 0.04914 | 0.01314 | 0.06869 | 0.00000 | 0.00000 |
| | Adults (18+ years-old) HIV+ ART-S | 0.11845 | 0.05563 | 0.00000 | 0.04233 | 0.00000 |
| | Adults (18+ years-old) HIV+ ART-L | 0.21324 | 0.06878 | 0.00000 | 0.00000 | 0.02797 |

[§] Pneumococcal conjugate vaccine (PCV13) serotypes (VT) and non-PCV13 serotypes (NVT)
[†] Human immunodeficiency virus (HIV) status in adults
[†] Antiretroviral therapy (ART)
[†] HIV- (without HIV infection), HIV+ ART-S (with HIV and shorter duration of ART use of <3 months), HIV+ ART-L (with HIV and longer duration of ART use of >1 year)

| Supplementary Table 2. Household and community pneumococcal forces of infections on each Age/HIV group and their estimates of posterior median and 95% credible intervals. | | |
|---|---|---|
| Parameter | VT carriage | NVT carriage |
| | Posterior median (95% credible intervals) | Posterior median (95% credible intervals) |
| **Absolute total force of infection for household (hh_FOI)** | | |
| Young children (0-5 years-old) | 0.106, (0.053-0.175) | 0.411, (0.193-0.684) |
| Older children (6-17 years-old) | 0.059, (0.023-0.117) | 0.314, (0.129-0.567) |
| Adults (18+ years-old) HIV- | 0.102, (0.041-0.203) | 0.144, (0.045-0.311) |
| Adults (18+ years-old) HIV+ ART-S | 0.031, (0.008-0.082) | 0.239, (0.083-0.503) |
| Adults (18+ years-old) HIV+ ART-L | 0.088, (0.032-0.170) | 0.337, (0.107-0.685) |
| **Absolute total force of infection for community (cc_FOI)** | | |
| Young children (0-5 years-old) | 0.041, (0.007-0.114) | 0.352, (0.142-0.667) |
| Older children (6-17 years-old) | 0.088, (0.039-0.160) | 0.261, (0.111-0.483) |
| Adults (18+ years-old) HIV- | 0.016, (0.001-0.067) | 0.079, (0.014-0.202) |
| Adults (18+ years-old) HIV+ ART-S | 0.019, (0.003-0.051) | 0.138, (0.040-0.295) |
| Adults (18+ years-old) HIV+ ART-L | 0.042, (0.006-0.124) | 0.229, (0.077-0.463) |
| **Absolute total force of infection (hh_FOI+cc_FOI)** | | |
| Young children (0-5 years-old) | 0.150, (0.0819-0.245) | 0.771, (0.491-1.140) |
| Older children (6-17 years-old) | 0.150, (0.0843-0.238) | 0.580, (0.369-0.886) |
| Adults (18+ years-old) HIV- | 0.124, (0.0561-0.232) | 0.230, (0.117-0.418) |
| Adults (18+ years-old) HIV+ ART-S | 0.052, (0.0195-0.110) | 0.386, (0.212-0.674) |
| Adults (18+ years-old) HIV+ ART-L | 0.135, (0.0632-0.241) | 0.573, (0.309-0.972) |
| **Household fraction of total force of infection (hh_FOI/[hh_FOI+cc_FOI])** | | |
| Young children (0-5 years-old) | 0.723, (0.421-0.945) | 0.537, (0.283-0.782) |
| Older children (6-17 years-old) | 0.403, (0.179-0.657) | 0.546, (0.265-0.787) |
| Adults (18+ years-old) HIV- | 0.866, (0.518-0.994) | 0.651, (0.254-0.929) |
| Adults (18+ years-old) HIV+ ART-S | 0.621, (0.231-0.921) | 0.636, (0.285-0.891) |
| Adults (18+ years-old) HIV+ ART-L | 0.674, (0.306-0.939) | 0.597, (0.255-0.854) |
| **Community fraction of total force of infection (cc_FOI/[hh_FOI+cc_FOI])** | | |
| Young children (0-5 years-old) | 0.277, (0.055 0.579) | 0.463, (0.218 0.717) |
| Older children (6-17 years-old) | 0.597, (0.343 0.821) | 0.454, (0.213 0.735) |
| Adults (18+ years-old) HIV- | 0.134, (0.006 0.482) | 0.349, (0.071 0.746) |
| Adults (18+ years-old) HIV+ ART-S | 0.379, (0.079 0.769) | 0.364, (0.109 0.715) |
| Adults (18+ years-old) HIV+ ART-L | 0.326, (0.061 0.694) | 0.403, (0.146 0.745) |

§ Pneumococcal conjugate vaccine (PCV13) serotypes (VT) and non-PCV13 serotypes (NVT)
† Human immunodeficiency virus (HIV) status in adults
† Antiretroviral therapy (ART)
† HIV- (without HIV infection), HIV+ ART-S (with HIV and shorter duration of ART use of <3 months), HIV+ ART-L (with HIV and longer duration of ART use of >1 year)

| Supplementary Table 3. Age/HIV stratified pneumococcal carriage duration, force of infection, and serotype competition relative risk, and their estimates of posterior median and 95% credible intervals. | | |
|---|---|---|
| Parameter | VT carriage | NVT carriage |
| | Posterior median (95% credible intervals) | Posterior median (95% credible intervals) |
| Carriage duration (weeks) | | |
| Young children (0-5 years-old) | 3.46, (1.38-16.4) | 5.00, (2.79-10.2) |
| Older children (6-17 years-old) | 1.46, (0.79-3.55) | 1.68, (1.05-2.83) |
| Adults (18+ years-old) HIV- | 1.58, (0.72-6.65) | 1.38, (0.78-2.89) |
| Adults (18+ years-old) HIV+ ART-S | 1.25, (0.62-4.32) | 1.21, (0.72-2.21) |
| Adults (18+ years-old) HIV+ ART-L | 3.65, (1.24-49.7) | 1.43, (0.82-2.75) |
| Serotype competition relative risk | | |
| NVT risk while carrying VT | - | 0.203, (0.020-0.560) |
| VT risk while carrying NVT | 0.325, (0.068-0.801) | - |

[§] Pneumococcal conjugate vaccine (PCV13) serotypes (VT) and non-PCV13 serotypes (NVT)
[†] Human immunodeficiency virus (HIV) status in adults
[†] Antiretroviral therapy (ART)
[†] HIV- (without HIV infection), HIV+ ART-S (with HIV and shorter duration of ART use of <3 months), HIV+ ART-L (with HIV and longer duration of ART use of >1 year)

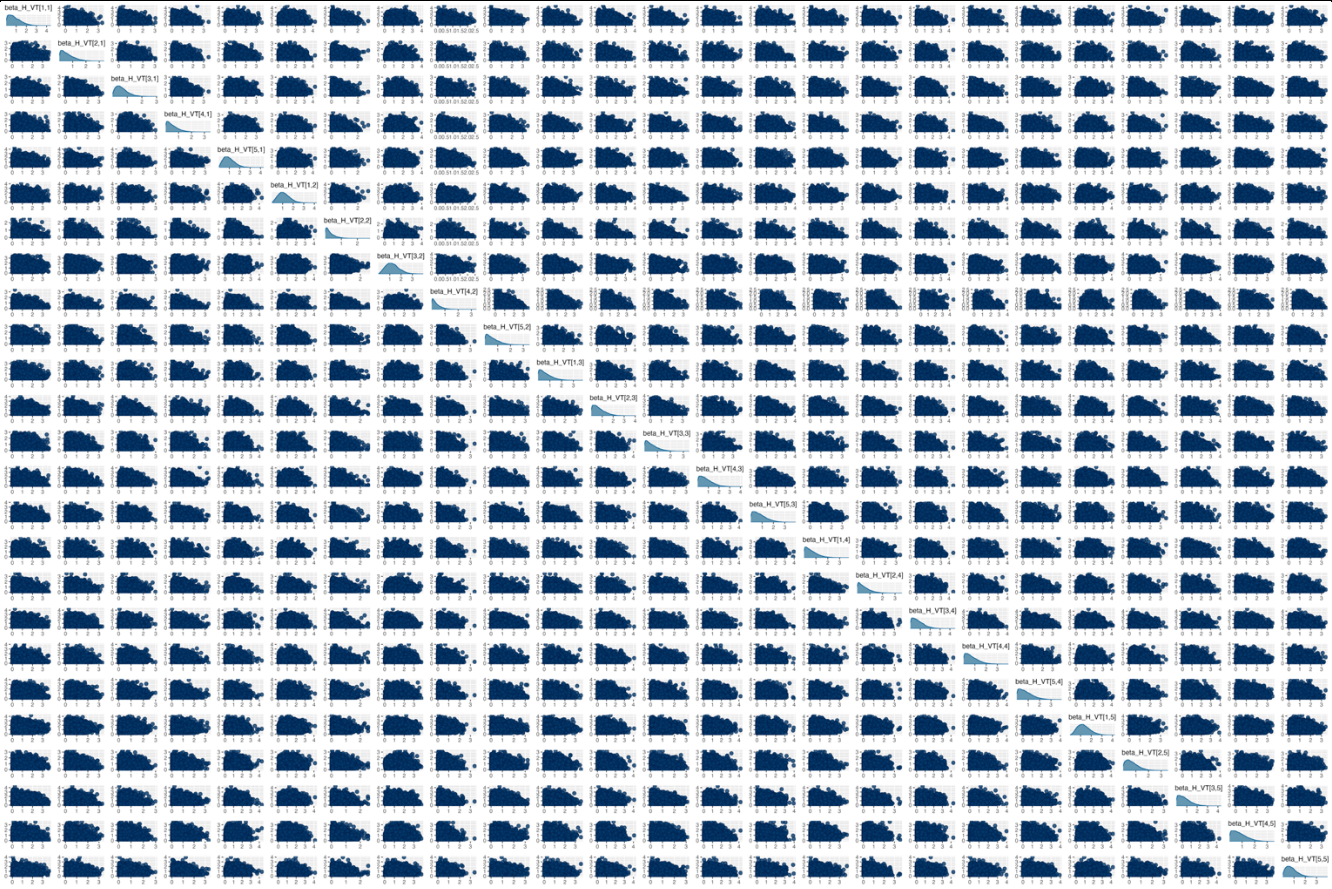

Supplementary Figure 1. Trace plots (hairy caterpillars) and posterior distribution (density plots) of (PCV13) vaccine serotype transmission parameters from 'who acquires infection from who' (WAIFW) matrices representing the transmission routes between different age/HIV groups. Each trace plot shows 4 Markov chains and a total of 4000 samples with 1000 warmup samples obtained after sampling from the posterior distribution using a Hamiltonian Monte Carlo (HMC) No U-Turn Sampler (NUTS). The posterior samples were obtained by fitting a household pneumococcal transmission model to longitudinal pneumococcal carriage swabbing data using a full Bayesian Framework.

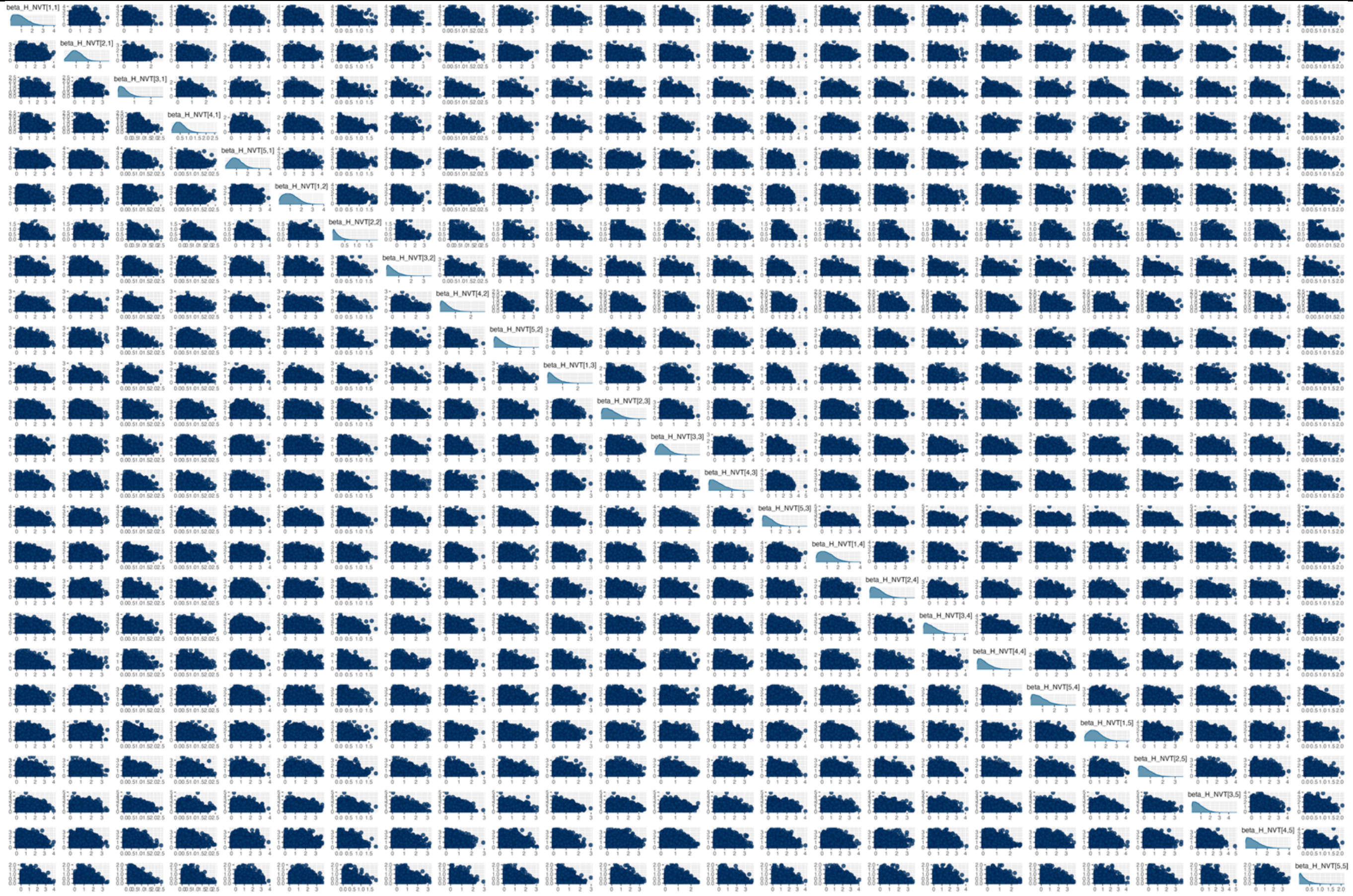

Supplementary Figure 2. Trace plots (hairy caterpillars) and posterior distribution (density plots) of (non-PCV13) non-vaccine serotype transmission parameters from ‘who acquires infection from who’ (WAIFW) matrices representing the transmission routes between different age/HIV groups. Each trace plot shows 4 Markov chains and a total of 4000 samples with 1000 warmup samples obtained after sampling from the posterior distribution using a Hamiltonian Monte Carlo (HMC) No U-Turn Sampler (NUTS). The posterior samples were obtained by fitting a household pneumococcal transmission model to longitudinal pneumococcal carriage swabbing data using a full Bayesian Framework.

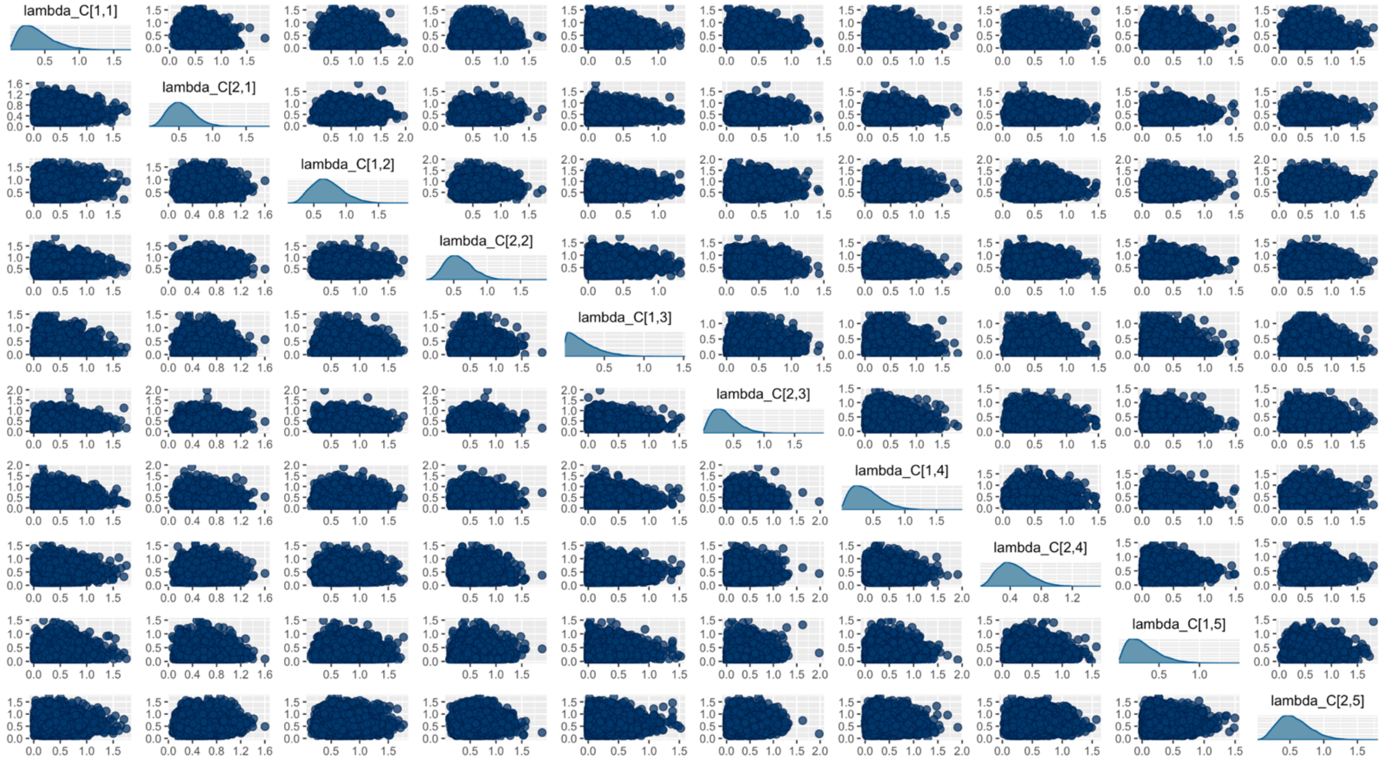

Supplementary Figure 3. Trace plots (hairy caterpillars) and posterior distribution (density plots) of (PCV13) vaccine serotype and non-vaccine serotype force of infection parameters of different age/HIV groups. Each trace plot shows 4 Markov chains and a total of 4000 samples with 1000 warmup samples obtained after sampling from the posterior distribution using a Hamiltonian Monte Carlo (HMC) No U-Turn Sampler (NUTS). The posterior samples were obtained by fitting a household pneumococcal transmission model to longitudinal pneumococcal carriage swabbing data using a full Bayesian Framework.

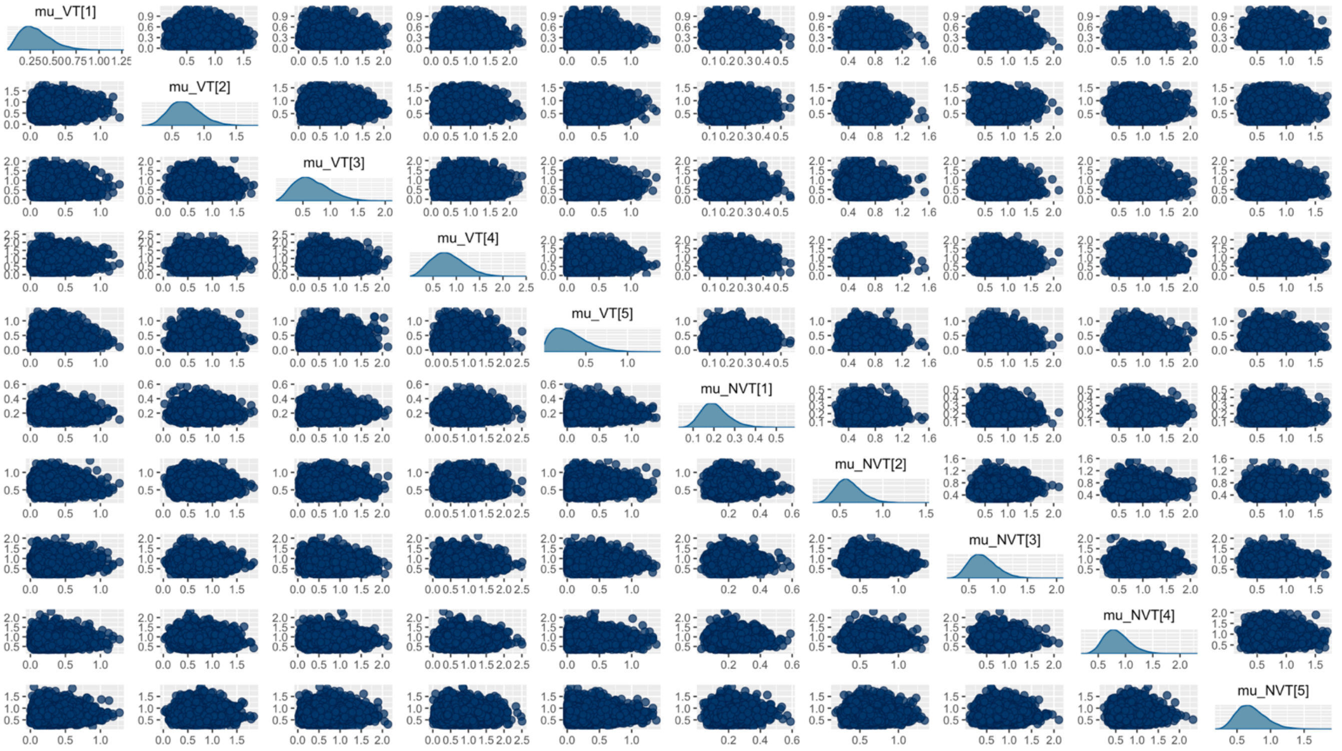

Supplementary Figure 4. Trace plots (hairy caterpillars) and posterior distribution (density plots) of (PCV13) vaccine serotype and non-vaccine serotype carriage clearance rate parameters of different age/HIV groups. Each trace plot shows 4 Markov chains and a total of 4000 samples with 1000 warmup samples obtained after sampling from the posterior distribution using a Hamiltonian Monte Carlo (HMC) No-U-Turn Sampler (NUTS). The posterior samples were obtained by fitting a household pneumococcal transmission model to longitudinal pneumococcal carriage swabbing data using a full Bayesian Framework.

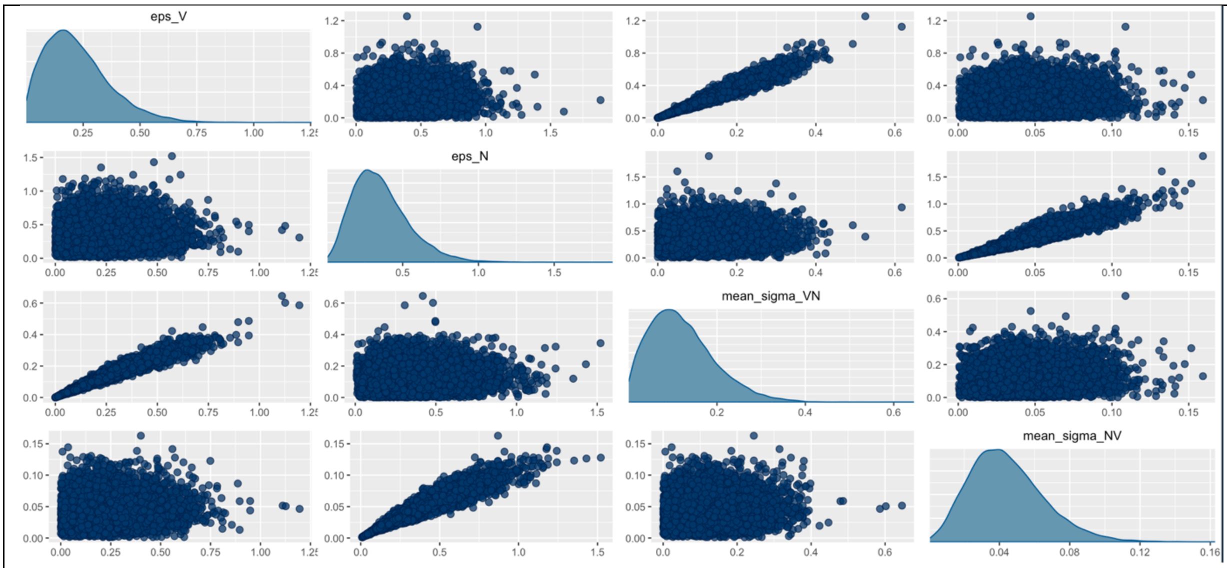

Supplementary Figure 5. Trace plots (hairy caterpillars) and posterior distribution (density plots) of serotype competition relative risk parameters. Each trace plot shows 4 Markov chains and a total of 4000 samples with 1000 warmup samples obtained after sampling from the posterior distribution using a Hamiltonian Monte Carlo (HMC) No-U-Turn Sampler (NUTS). The posterior samples were obtained by fitting a household pneumococcal transmission model to longitudinal pneumococcal carriage swabbing data using a full Bayesian Framework.

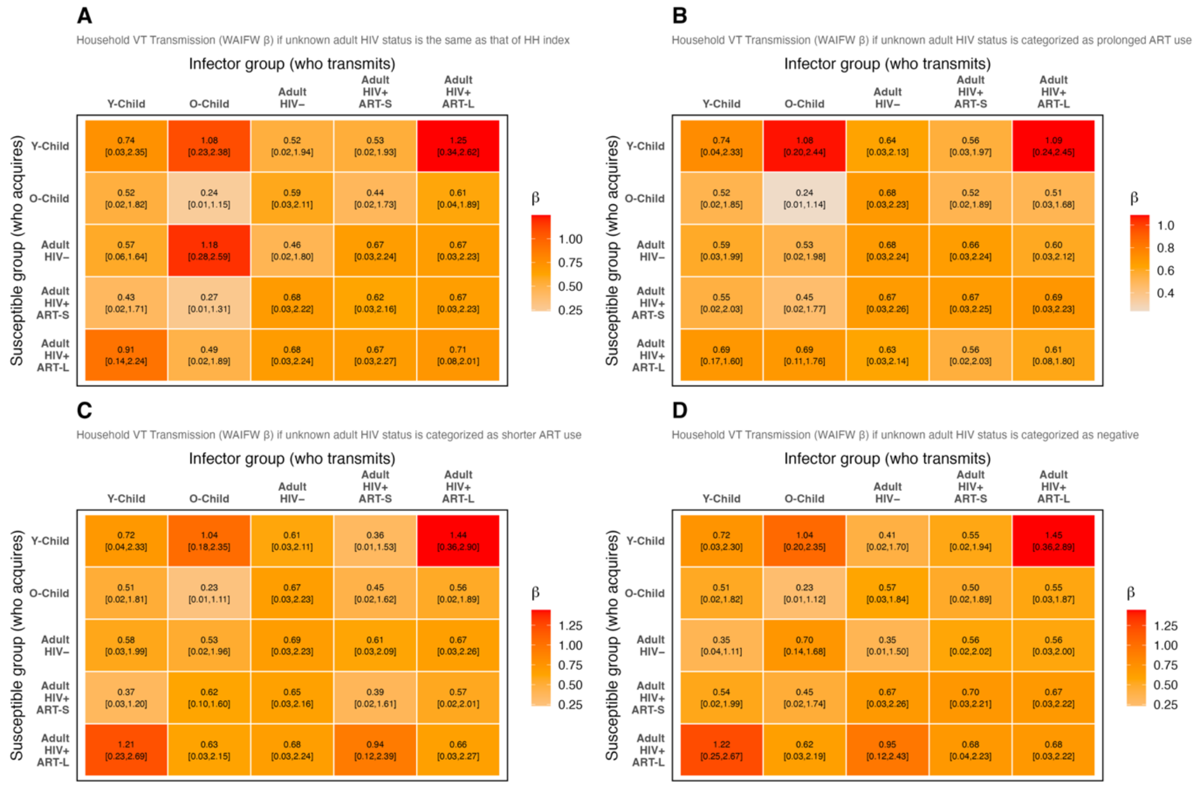

Supplementary Figure 6. Sensitivity analysis of who acquires infection (**vaccine serotypes**) from who (WAIFW) based on different categorisation of unknown data on adult human immunodeficiency virus (HIV) infection status. Number in each cell show posterior median and 95% credible intervals (CrI) of transmission events per week as estimated by fitting a household pneumococcal transmission model to longitudinal pneumococcal swabbing data and sampling from the posterior distribution using Hamiltonian Monte Carlo (HMC) No-U-Turn Sampler. The heatmaps show transmission rates from age/HIV group to another age/HIV group assuming that: (A) unknown household adult HIV status is the same as that of the household index with known HIV status, (B) unknown household adult HIV status is categorised as adults with HIV and prolonged duration (>1 year) of antiretroviral (ART) use, (C) unknown household adult HIV status is categorised as adults with HIV and shorter duration (<3 months) of ART use, and (D) unknown household adult HIV status is categorised as adults without HIV.

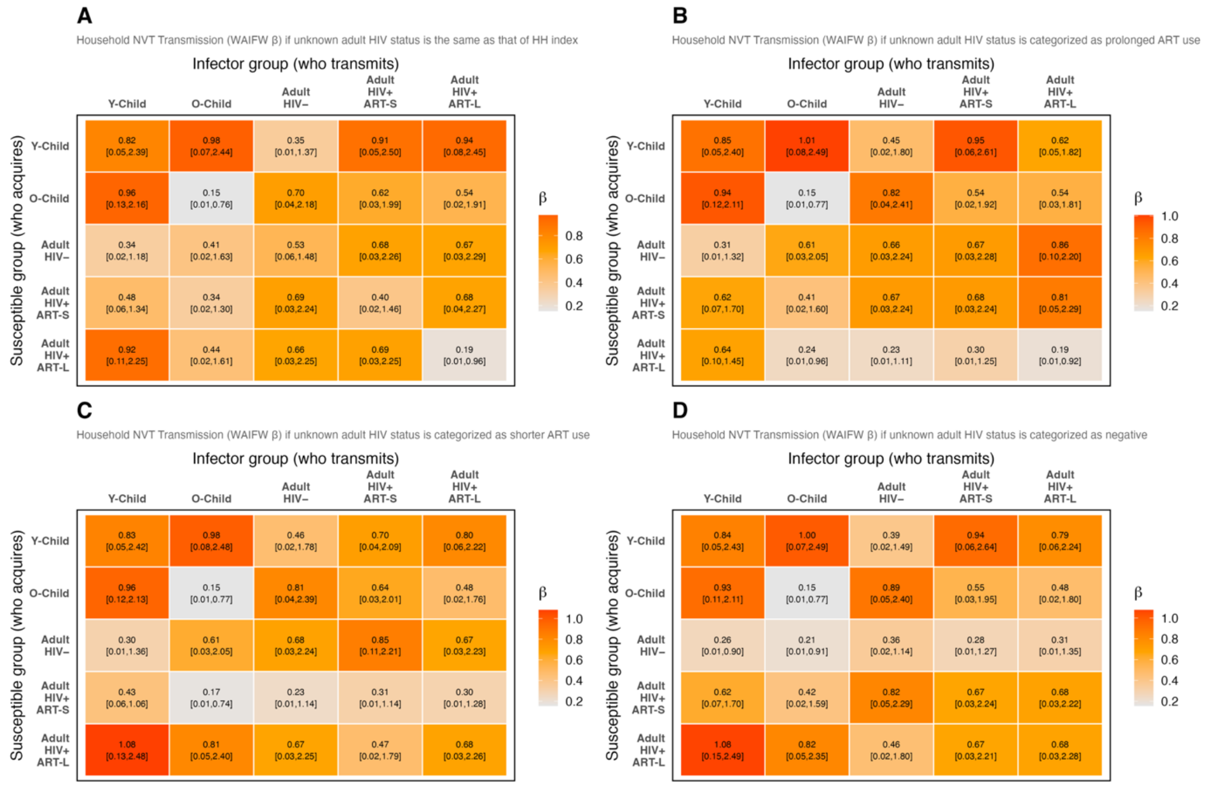

Supplementary Figure 7. Sensitivity analysis of who acquires infection (**non-vaccine serotypes**) from who (WAIFW) based on different categorisation of unknown data on adult human immunodeficiency virus (HIV) infection status. Number in each cell show posterior median and 95% credible intervals (CrI) of transmission events per week as estimated by fitting a household pneumococcal transmission model to longitudinal pneumococcal swabbing data and sampling from the posterior distribution using Hamiltonian Monte Carlo (HMC) No-U-Turn Sampler. The heatmaps show transmission rates from age/HIV group to another age/HIV group assuming that: (A) unknown household adult HIV status is the same as that of the household index with known HIV status, (B) unknown household adult HIV status is categorised as adults with HIV and prolonged duration (>1 year) of antiretroviral (ART) use, (C) unknown household adult HIV status is categorised as adults with HIV and shorter duration (<3 months) of ART use, and (D) unknown household adult HIV status is categorised as adults without HIV.

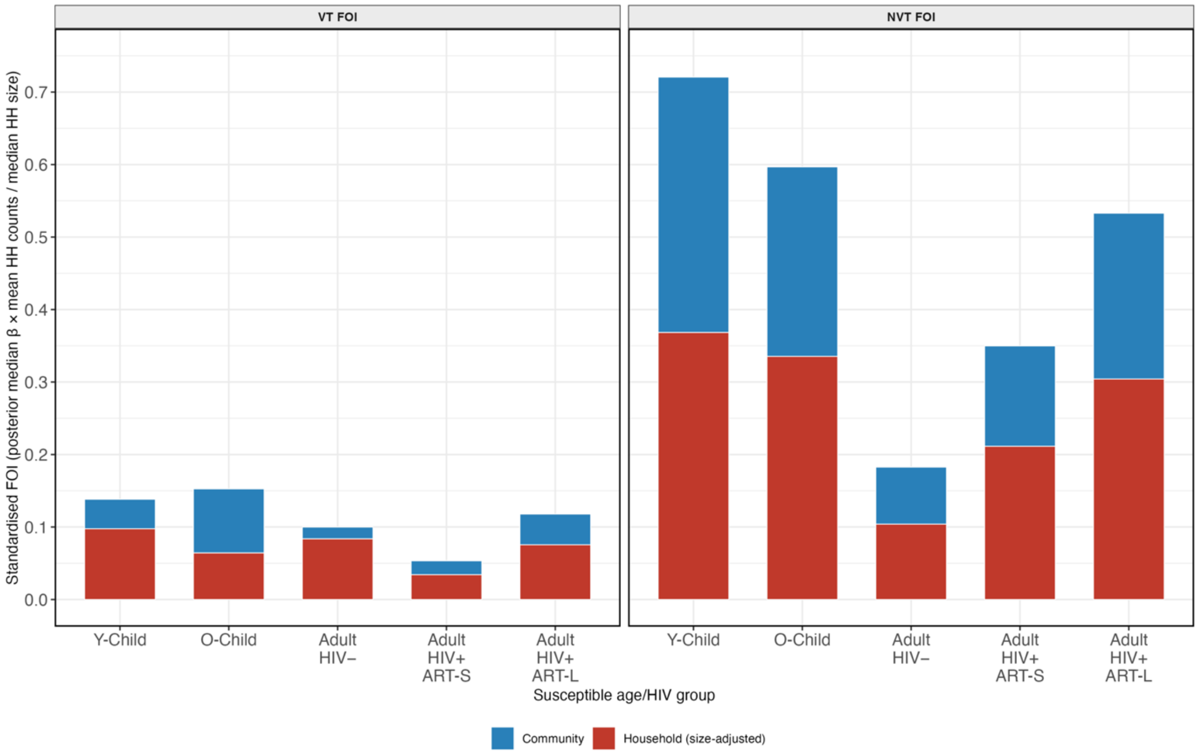

Supplementary Figure 8. Sensitivity analysis of the effect of standardized force of infection (FOI) on household vs community share of FOI. The FOI was standardized to median household size of 4 which removed the individual variation in household sizes (multiplying the posterior median transmission rate by mean household count and dividing by median household counts) and highlighted the structural household vs community contributions. The height of the bar refers to posterior median estimate of each age/HIV group stratified by household vs community source of FOI.

**Supplementary Text 1. Details of the household transmission modelling**

***Pneumococcal dynamic model.***

We develop a model that is a simplification of the complex dynamics of pneumococcal transmission. With the SIS model structure, an individual $i$ in age group $\alpha_i$ can move from being uncolonized ($S_i$) to become a VT carrier ($I_{i,VT}$) or NVT carrier ($I_{i,NVT}$) after experiencing age/HIV- and serotype group-specific FOI ($\lambda_{i,VT}$) or ($\lambda_{i,NVT}$), respectively. An individual can move back from a carrier state to uncolonized state over a time interval ($\delta t$) after clearing VT carriage ($\mu_{i,VT}$) or NVT carriage ($\mu_{i,NVT}$). VT carriers have a 'change of serotype' rate of $\phi_{i,VN,t} = \varepsilon_{VT} \times \lambda_{i,NVT,t}$ to switch to become NVT carriers without a clearance episode where $\varepsilon_{VT}$ is the relative risk to acquire NVT while carrying VT compared to uncolonized state. Likewise, NVT carriers have a 'change of serotype' rate of $\phi_{i,NV,t} = \varepsilon_{NVT} \times \lambda_{i,VT,t}$ to switch to become VT carriers without a clearance episode where $\varepsilon_{NVT}$ is the relative risk to acquire VT while carrying NVT compared to uncolonized state. The following set of equations describes the model probabilities of an individual in age/HIV group making transitions between the states in a short-time interval $\delta t$:

$$P_i(S \rightarrow I_{VT})_{\delta t} = \lambda_{i,VT} \times \delta t \qquad (1)$$

$$P_i(S \rightarrow I_{NVT})_{\delta t} = \lambda_{i,NVT} \times \delta t \qquad (2)$$

$$P_i(I_{VT} \rightarrow S)_{\delta t} = \mu_{i,VT} \times \delta t \qquad (3)$$

$$P_i(I_{NVT} \rightarrow S)_{\delta t} = \mu_{i,NVT} \times \delta t \qquad (4)$$

$$P_i(I_{VT} \rightarrow I_{NVT})_{\delta t} = \varepsilon_{VT} \times \lambda_{i,NVT} \times \delta t \qquad (5)$$

$$P_i(I_{NVT} \rightarrow I_{VT})_{\delta t} = \varepsilon_{NVT} \times \lambda_{i,VT} \times \delta t \qquad (6)$$

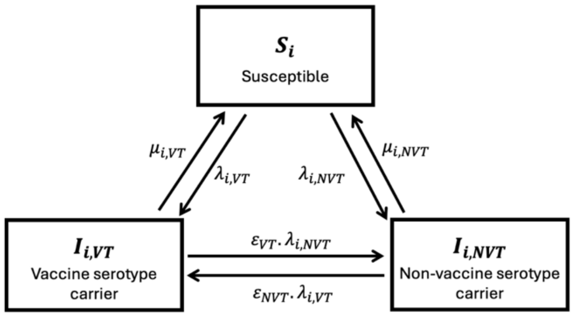

The Susceptible-Infected-Susceptible (SIS) model structure that captures the dynamics of pneumococcal household carriage in Blantyre, Malawi. This model is used to quantify within-household pneumococcal transmission routes from the source (infector) to the target (susceptible) through who acquires infection from who (WAIFW) matrix parameterisation. The model is also used to quantify household and community (background) forces of infection after a 15-year longstanding routine pneumococcal congugate vaccine (PCV13) program. The model states $S, I_{VT}, I_{NVT}$, refer to uncolonized, colonized with PCV13 serotypes (VT), colonized with non-PCV13 serotypes (NVT). Parameters $\mu$ refer to carriage clearance rate, $\lambda$ refer to the force of infection, and $\varepsilon$ refer to relative risk of acquiring a competitor serotype while carrying a resident serotype compared to uncolonized state. The subscripts $VT$ refer to vaccine serotypes, $NVT$ to non-PCV13 serotypes, $i$ to each of the five age exposure groups (younger child, older child, adult without human immunodeficiency syndrom (HIV), adults with short-term ART, adults with long-term ART).

***Model likelihood.***

Let $X_{i,t} \in \{1, 2, 3\}$ denote the observed carriage state of individual $i$ at visit $t$, and let $\alpha_i \in \{1, \ldots, 5\}$ be their age/HIV group. Let $H$ denote the household of individual $i$. At a transition $t - (t+1)$ for $t = (1, 2)$, the total FOI for serotype type $k \in \{\text{VT, NVT}\}$ acting on individual $i$ comprised a household component and a community component:

$$\lambda_{i,k,t} = \left(\sum_{b=1}^{5} \beta_k^H[\alpha_i, b] \times \left[I_{i,k,b,t}^H \times (N_t^H - 1)^{-w}\right]\right) + \left(\lambda_k^C[\alpha_i] \times \pi_{k,\alpha_i,t}\right) \quad (7)$$

Where $\beta_k^H[\alpha_i, b]$ is the household WAIFW transmission rate from infectors of age/HIV group $b$ to susceptibles of age/HIV group $\alpha_i$ for serotype group $k$ per week. $I_{i,k,b,t}^H$ is the count of the serotype group-$k$ carriers of age/HIV group $b$ in household $H$ at visit $t$, excluding the individual $i$ themselves. $(N_t^H - 1)^{-w}$ is the total number of other household members at visit $t$, where $w$ is the density correlation factor controlling the extent of density-dependency. $\lambda_k^C[\alpha_i]$ is the community (background) FOI for age/HIV group $\alpha_i$ and serotype group $k$. $\pi_{k,\alpha_i,t}$ is the observed cross-sectional community prevalence of serotype group $k$ in age/HIV group $\alpha_i$ at visit $t$ that scales the community FOI. We assumed the density scaling factor of $w$=1 based on previous studies [1, 2], such that the forces of infection are proportional to the fraction of household members who are carriers.

Thus, the instantaneous rate matrix $Q$ for an individual $i$ of exposure group $\alpha_i$ with the forces of infection $\lambda_{i,VT}$ and $\lambda_{i,NVT}$ was a 3 × 3 valid matrix with rows summing to zero and the $jth$ diagonal element satisfying $Q[j,j] = -\sum_{l \neq j} Q[j,l]$.

$$Q_{i,t} = \begin{pmatrix} -(\lambda_{i,VT,t} + \lambda_{i,NVT,t}) & \lambda_{i,VT,t} & \lambda_{i,NVT,t} \\ \mu_{i,VT}[\alpha_i] & -(\mu_{i,VT}[\alpha_i] + \phi_{i,VN,t}) & \phi_{i,VN,t} \\ \mu_{i,NVT}[\alpha_i] & \phi_{i,NV,t} & -(\mu_{i,NVT}[\alpha_i] + \phi_{i,NV,t}) \end{pmatrix} \quad (8)$$

Likewise, the transition probability matrix $P$ over one week ($\delta t$ = 1) was obtained by matrix exponentiation, $P_{i,t} = \exp\,(Q_{i,t} \times \delta t)$. The element $P_{i,t}[s, s']$ gives the probability of transitioning from carriage state $s$ to state $s'$ for individual $i$ between visits $t$ and $t + 1$. The matrix exponential was computed exactly for each individual-transition pair using Stan's 'matrix_exp()' function, which applies the scaling-and-squaring algorithm, as alternative to numerical ODE integration, and produces exact finite-time transition probabilities for any parameter configuration.

Assuming the conditional independence of transitions across individuals and across time given the observed household composition, the full log-likelihood of data given the parameter vector $\theta$ is given below, with $\theta \in \{\beta_k^H[\alpha_i, b],\, \lambda_k^C[\alpha_i],\, \mu_{i,k},\, \varepsilon_k\}$ and $X$ as all observed carriage state sequences. A lower bound (10e-12) was imposed on individual transition probabilities within the log argument to prevent numerical underflow in regions of the parameter space where the matrix exponential produces near-zero entries due to extreme parameter values during warm-up.

$$\ell(X \mid \theta) = \sum_{i=1}^{302} \sum_{t=1}^{2} log\ (P_{i,t}[X_{i,t}, X_{i,t+1}]) \qquad (9)$$

***Model parameterisation.***

*Household WAIFW transmission matrices*. The matrices, $\beta_{VT}^H$ and $\beta_{NVT}^H$, parameterise within-household pneumococcal transmission for VT and NVT, respectively. Matrix element $\beta_k^H[a, b]$ represents the number of serotype group $k$ carriage events a susceptible individual in age/HIV group $\alpha$ acquires from a carrier in age/HIV group $b$ within household per week. Thus, directed routes per serotype group for child-to-child, child-to-adult, adult-to-child, and adult-to-adult combinations across the two child age groups and three adult HIV strata were estimated.

*Community forces of infection*. The matrix $\lambda_k^C$ estimates serotype group-specific community acquisitions. The matrix element $\lambda_k^C[\alpha]$ multiplied the observed community prevalence $\pi_{k,\alpha,t}$ at visit $t$, as the proportion of individuals of age/HIV group $\alpha$ with $k$ carriage across all households, allowing the community FOI to vary over time without requiring additional parameters.

*Clearance rates*. Parameters, $\mu_{VT}[\alpha]$ and $\mu_{NVT}[\alpha]$, were the instantaneous rate at which carriers of each age/HIV group $\alpha$ cleared VT and NVT carriage, respectively. The mean carriage duration for age/HIV group $\alpha$ was $1/\mu_k[\alpha]$ weeks (exponential distribution assumption). The distinct age/HIV clearance rates captures the heterogeneity in immune clearance of pneumococci.

*Serotype competition*. Parameters, $\varepsilon_{VT}$ and $\varepsilon_{VT}$, estimated the relative risk of NVT and VT colonisation while already carrying VT and NT, respectively, compared with a fully susceptible individual. A value of of $\varepsilon$=0 implied complete exclusion where a resident serotype always exclude competitors, 0<$\varepsilon$<1 competitive exclusion with resident serotype allowing some level of co-colonisation, $\varepsilon$=1 resident serotype fully susceptible to competitors, and $\varepsilon$>1 resident serotype facilitating acquisition of a competitor. A relative competition was calculated as $\varepsilon_{VT}/\varepsilon_{NVT}$ with a value >1 implying that NVT displaces VT more readily per unit of exposure pressure.

*Derived parameters*. We derived the rate of serotype group switching and the decomposed FOI. The rates of serotype group switching between VT and NVT were computed at each transition by multiplying serotype group relative risk to co-colonisation with the FOI and keeping its posterior distributions with full uncertainty propagation. Similarly, for every posterior draw, for every individual, and for both transitions, we decomposed the total FOI into household and community sources as in equation 7. The estimates of the forces of infection were accumulated within each age/HIV group, averaged over individuals and transitions, and kept as posterior distributions with full uncertainty propagation. This enabled estimation of the absolute and share of FOI to each source.

**Parameter inference**

All model parameters were jointly estimated within a fully Bayesian framework. The log-posterior distribution of parameters given the data was given as: $log\ P(\theta \mid X) \propto \ell(X \mid \theta) + log\ p(\theta)$, where $\ell(X \mid \theta)$ is the log-likelihood of data given the parameter vector $\theta$ expressed in equation 9, $\mathrm{p}(\theta)$ the prior distribution of parameters, and $X$ denotes all observed carriage state sequences. Posterior inference was performed using Hamiltonian Monte Carlo (HMC) with the No-U-Turn Sampler (NUTS) [3], as implemented in Stan v2.38 [4], and accessed via the RStan interface [5]. Weakly informative prior distributions were specified as described in Table 1.

Sampling was performed with 4 independent Markov chains, each initialised from random starting values, with 1,000 warmup (adaptation) iterations and 5,000 posterior sampling iterations, resulting in 16,000 post-warmup posterior draws. The NUTS target acceptance rate was set to 0.95 with maximum leapfrog tree depth of 12. Convergence was assessed using a scale reduction factor (target: *Ṛhat* <1.01) and both bulk and tail effective sample sizes (target: ESS ≥1000 per chain). HMC-specific diagnostics including divergent transitions (target: zero post-warmup) and energy Bayesian fraction of missing information (target: E-BFMI ≥ 0.2) were assessed. Posterior predictive checks compared the observed proportions of individuals in each carriage state at visits 2 and 3 with replicated samples drawn from the posterior predictive distribution: Parameter summaries were reported as posterior medians with 50% and/or 95% credible intervals (95%CrI). All the computation was done using R software pakage (RStudio v2024.04.0, R v4.5.3) [6]. The code is freely and publicly available under the MIT License and can be accessed at: https://github.com/deusthindwa/spn.hhtransmission.dynamic.model.malawi.

| The prior distribution or fixed value for each parameter used during model fitting | | | |
|---|---|---|---|
| Parameter | Prior distribution | Source | Rationale |
| $\beta^{H}_{VT}[a,b]$<br>$\beta^{H}_{NVT}[a,b]$ | Half-Normal(0, 1) | Althouse ([7]) | Truncated positive constraint for within-household transmission per week. |
| $\lambda^{C}_{VT}[a]$<br>$\lambda^{C}_{NVT}[a]$ | Half-Normal(0, 0.5) | Thindwa et al. ([8])<br>Althouse ([7]) | Truncated positive constraint for community exposure expected smaller than household. A narrower scale provides additional shrinkage. |
| $\mu_{VT}[a]$<br>$\mu_{NVT}[a]$ | Normal(1, 0.5) × (0, ∞) | Phiri et al. ([9])<br>Thindwa et al. ([8]) | Centred on 1 clearance event per week. Shared hyperparameters pool information across groups to correct for small sample sizes. |
| $\varepsilon_{VT}$<br>$\varepsilon_{NVT}$ | Exponential(2) | Kamng'ona et al. [10] | Mean 0.5 (prior favours competitive exclusion, $\varepsilon$ <1, but allows $\varepsilon$ >1 if supported by the data) |
| $w$ | Fixed, $w$=1 | Melegaro et al. ([1]) | A density correlation factor scaling the extent of density-dependency within household |
